# Formulation of a spatiotemporal model for analysis of neonatal mortality amidst SDGs intervention. The case of Uganda

**DOI:** 10.1101/2025.04.17.25326005

**Authors:** Bamwebaze George, Gichuhi A. Waititu, Richard O Awichi, Atinuke Olusola Adebanji

## Abstract

The study aimed to formulate a dynamic linear model within a Bayesian framework to conduct a spatiotemporal analysis of neonatal mortality in Uganda during the SDGs intervention. The study ably formulated the model based on appropriate health-related covariates while considering the spatial and temporal dimensions of the data whose variable of interest (dependent variable) was a quantitative variable measuring the monthly rates of neonatal mortality (number of newborns dying within their first 28 days of life) at the district level of the country. Through Markov Chain Monte Carlo (MCMC) simulations, the study was able to assess the applicability of the model based on simulated data covering 14 years starting from January 2010. Using a Bayesian approach through the Kalman filtering technique, the study estimated the parameters of the formulated model. The study used the same technique through Gibbs sampling to extract meaningful information from the simulated data and provide reliable forecasts for the rates of neonatal mortality.

## Introduction

The burden of Neonatal mortality has continued to rise the mortality rates of most countries [2]. A healthy economy with a minimal mortality rate is a desire for every country worldwide. Neonatal (newborn) mortality is defined for this study as death within the first 28 days of life per 1000 live births as established by the World Health Organization [1]. As noted by [2], reducing neonatal mortality is an essential part of SDG 3, section 3.2: *countries should aim to reduce neonatal mortality to at least 12 per 1*,*000 live births and under-five mortality to at least 25 per 1*,*000 live births by 2030* and achieving this requires an understanding of the levels of and trends in neonatal mortality. According to a study by [3], 5.2 million children died before reaching their fifth birthday in 2019 with almost half of those deaths, 2.4 million occurring in the first month of life despite the countries’ efforts to suppress the death rates for instance UN member states’ intervention of coming up with the MDGs and SDGs.

In Uganda like on the global scene, most studies such as [4] [5] [6] on neonatal mortality have put their focus on risk factors ignoring measuring the progress and forecasting the situation given the fact that the targeted 2030 to achieve the SDGs is fast approaching. More so, these studies did not take into account time and space influence. This study aimed at formulating a dynamic linear model to analyze the situation of Neonatal mortality using a case study of Uganda to simultaneously investigate its persistent patterns over time and space and illuminate any unusual patterns. As mentioned by [9], majority of time series models including the well known ones like autoregressive (AR), moving average (MA), autoregressive integrated moving average (ARIMA), and autoregressive moving average (ARMA), are helpful for handling stationary data, otherwise they get limited. This is further supported by 10, [11], [12], [13], [14] as they suggest that state space models offer a very rich class of models that have several advantages. For example, they do not require stationarity, which eliminates the need to transform the data since data transformation leads to loss of some important components in the data which at times leads to less accurate results. In order to formulate a Dynamic Linear Model, the study simulated data for health care related factors and health care policies that were put in place by the country as a way of achieving SDG 3.2. The health policies based on are as shown in (1).

## Materials and Methods

### Data Types and Sources

To acheive the study objective, simulated data basing on appropriated distributions was used. Monthly data on the the following variables was simulated for a period of 14 years stating from January 2010. A case study of Uganda was used. Uganda is a landlocked country in eastern part of Africa.

### Study Variables

The dependent variable was Neonatal measured as the monthly number of newborn children dying within their first 28 days of life per district obtained by summing neonatal deaths within 0-7 and 8-28 first days of life

The independent variables (covariates) were;

1. Cost measured as the monthly amount of money the district receives from Ministry Of Finance through the Ministry Of Health for the implementation of the seven policies obtained by dividing the annual amount by 12.
2. Year measured as the calendar year for which the data was recorded.
3. Month measured as the month of the calendar year for which the data was recorded.
4. District measured as name of the district from which the data was recorded.
5. Region measured as the main administrative region in which the district is located since this data is reported monthly from districts not regions. Therefore regrouping of districts will be done to attain regions during Data cleaning and Editing. The regions will be coded as 1 for Central, 2 for Western, 3 for Eastern and 4 for Northern.
6. HealthfacDensity measured as the number of health centres in the district.
7. HcareAccess measured as number of government healthcare providers in the district.
8. HcenterAccess measured as the distance in meters to the nearest health center from the furthest household in the district.
9. MaternalEduc measured as average number of antenatal visits by women in the district per month.
10. HholdInc measured as the average monthly household income in the district.
11. Season measured as the changes in weather conditions in the country captured as (1 Dry, 0 Wet).
12. SDGintro measured as the time of the start of SDGs during the observation period captured as (1 After, 0 Before).
13. HcarepolicyInfo measured as the time of the start of the 7 healthcare policies shown in Table 1 during the observation period captured as (1 After, 0 Before).
14. Observ measured as observation time running from 1 in January 2010 to 168 in December 2023.

**Table 1.**
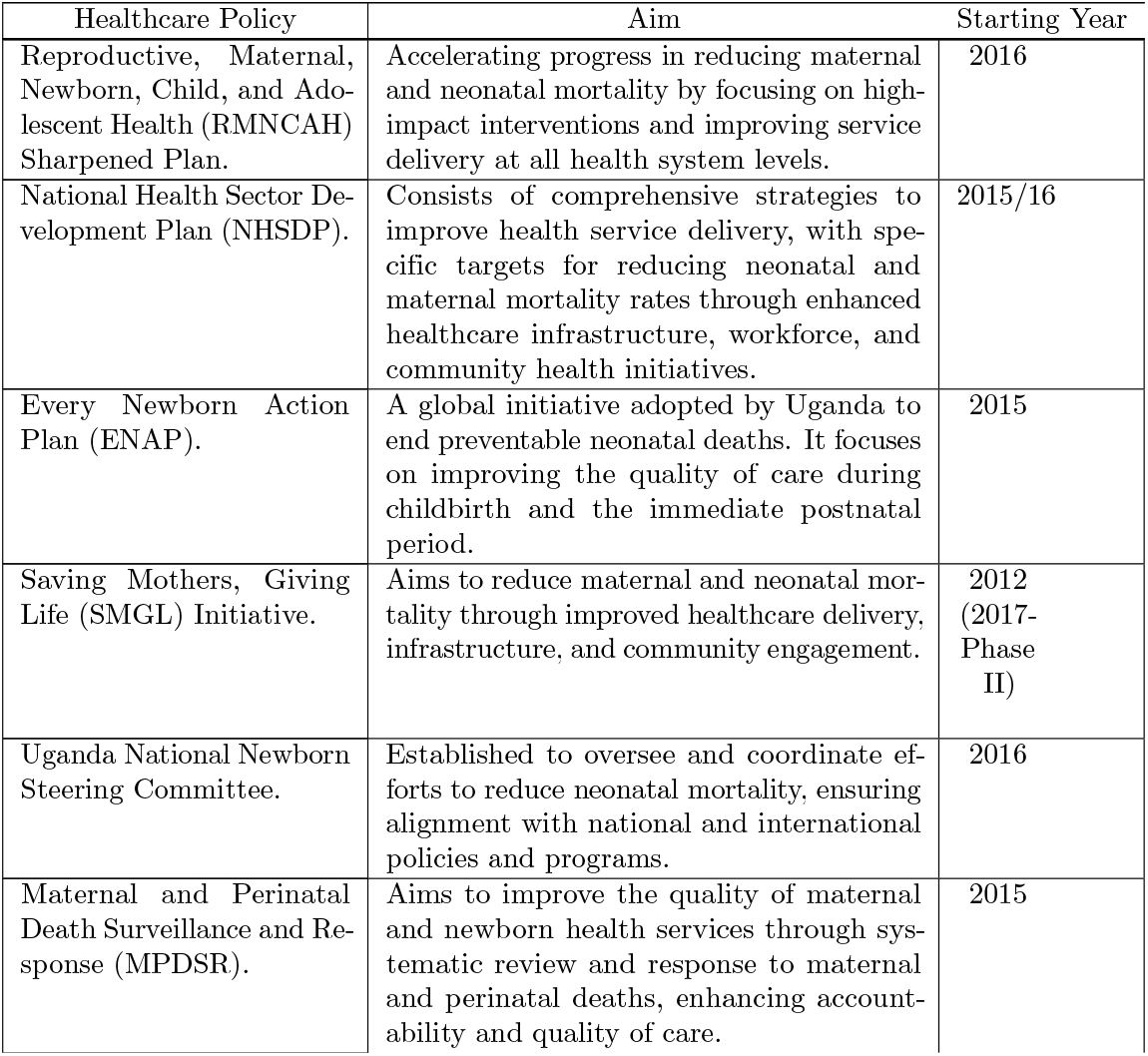
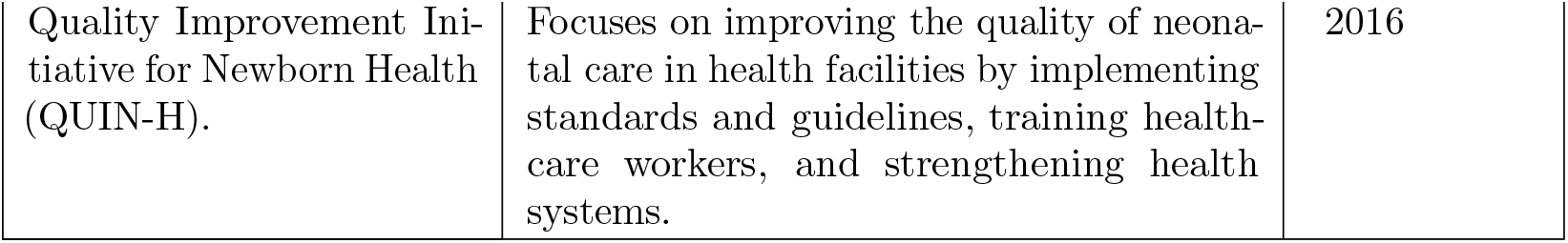
Policies in place to reduce neonatal mortality in Uganda.

| Healthcare Policy | Aim | Starting Year |
| --- | --- | --- |
| Reproductive, Maternal, Newborn, Child, and Adolescent Health (RMNCAH) Sharpened Plan. | Accelerating progress in reducing maternal and neonatal mortality by focusing on high-impact interventions and improving service delivery at all health system levels. | 2016 |
| National Health Sector Development Plan (NHSDP). | Consists of comprehensive strategies to improve health service delivery, with specific targets for reducing neonatal and maternal mortality rates through enhanced healthcare infrastructure, workforce, and community health initiatives. | 2015/16 |
| Every Newborn Action Plan (ENAP). | A global initiative adopted by Uganda to end preventable neonatal deaths. It focuses on improving the quality of care during childbirth and the immediate postnatal period. | 2015 |
| Saving Mothers, Giving Life (SMGL) Initiative. | Aims to reduce maternal and neonatal mortality through improved healthcare delivery, infrastructure, and community engagement. | 2012<br>(2017-Phase II) |
| Uganda National Newborn Steering Committee. | Established to oversee and coordinate efforts to reduce neonatal mortality, ensuring alignment with national and international policies and programs. | 2016 |
| Maternal and Perinatal Death Surveillance and Response (MPDSR). | Aims to improve the quality of maternal and newborn health services through systematic review and response to maternal and perinatal deaths, enhancing accountability and quality of care. | 2015 |
| Quality Improvement Initiative for Newborn Health (QUIN-H). | Focuses on improving the quality of neonatal care in health facilities by implementing standards and guidelines, training health-care workers, and strengthening health systems. | 2016 |

### Nature of and Rationale for the Model

The study opted for a dynamic linear model to monitor the impact of the SDG intervention on neonatal mortality by assessing the situation of neonatal mortality before and after its introduction. The reason for adopting a dynamic linear model is the fact that dynamic linear models constitute a flexible class of models that can deal with typical features in policy monitoring and data at large.

### General form of a Dynamic Linear Model

As a way of achieving the study objective: formulation of a Dynamic linear model, a general Gaussian dynamic linear model was based on, presented as follows:

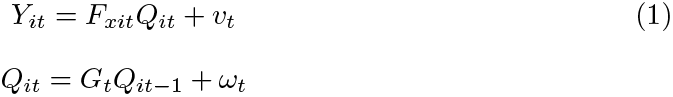

where *Y*_*it*_ represents observation for the *i*^*th*^ state at time t.

*F*_*xit*_ denotes a function of the m covariates for the *i*^*th*^ state at time t specified as

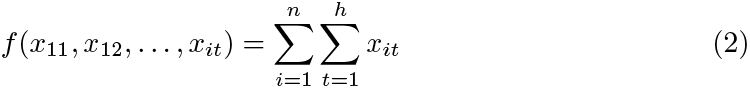

*Q*_*it*_ is a vector of time carrying parameters

*G*_*t*_ is a transition matrix depicting how the model parameters evolve over time.

*v*_*t*_ ∼𝒩 (0, *V*_*i*,*t*_) and *ω*_*t*_ ∼𝒩 (0, *W*_*i*,*t*_) are respectively the observation and evolution errors

### The Formulated Study Model

The model has been formulated as follows;

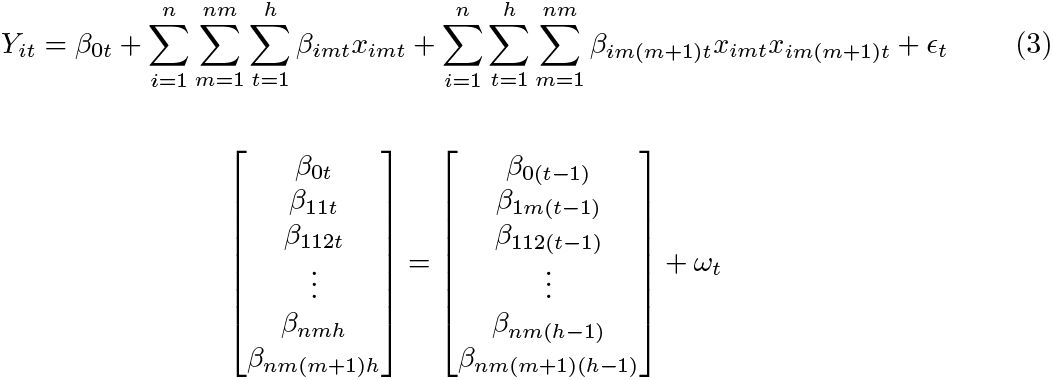

Where:

*Y*_*it*_ represents the *i*^*th*^ district neonatal mortality rate at time t.

*x*_11_, *x*_12_, …, *x*_*nh*_ are the m covariates including the maternal healthcare policies and interventions in which case a code 0 is used for Before and 1 for After the introduction of a given policy/intervention during the observation time for a given policy or intervention.

*x*_*itm*_*x*_*itm*(*m*+1)_ is the interaction covariate between the *m*^*th*^ *and* (*m* + 1)^*th*^ covariate including the maternal healthcare policies and interventions with *β*_*itm*(*m−*1)_ as the their respective coefficients,

*β*_0*t*_ denotes the time varying intercept.

*β*_111_, *β*_121_, …, *β*_*nhm*_ are the time varying coefficient associated with each of the m covariates including the maternal healthcare policies and interventions.

*ϵ*_*t*_ and *ω*_*t*_ are the observation and evolution error terms respectively where *ϵ*_*t*_ ∼ 𝒩 (0, *V*_*t*_) and *ω*_*t*_ ∼ 𝒩 (0, *W*_*t*_).

### Model Assumptions

The following properties will be checked for:

Auto-correlation:

The Residuals should not be autocorrelated.

Homoscedasciticity:

The residuals should have constant Variance.

Normality:

The residuals are expected to be normally distributed.

### Parameter Estimation

Since the study had to make a comparison of the neonatal mortality situation in the country before and after the introduction of SDGs implying that past values had much influence on future values for this study, the Bayesian approach using the Kalman filtering technique was considered the best method for estimating study model parameters considering the fact that Bayesian approach interests itself in how a particular part of the data depends on the other data parts.

Considering the fact that Bayesian inference has its roots from Bayes rule as noted in the studies by [7] and [8], to get the final parameter estimates, the study made use of the theorem through the posterior density function basing on;

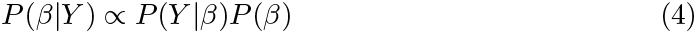

where

*β* represents a vector of parameters to be estimated

Y represents the observed data which is the district’s monthly neonatal mortality. P(*β*) represents the probability distribution of the parameter vector being estimated, that is to say, the update in the light of new data. This constitutes the prior probability.

*P* (*Y*|*β*) is the probability distribution of the data given the parameter being estimated which constitutes the likelihood.

*P* (*β*|*Y*) is the probability distribution of the parameter vector being estimated given the data Y. This constitutes the posterior probability of the estimated parameter (updated upon new data).

### Procedure followed to estimate the model parameters

Rewriting the formulated model presented in (3) which is expressed in the form of two equations; observation and state equations, bearing in mind the fact that the process of bayesian inference involves passing from a prior distribution, *P* (*β*) to a posterior distribution, *P* (*β*|*Y*), estimation of its parameters was done through the Kalman filtering technique.

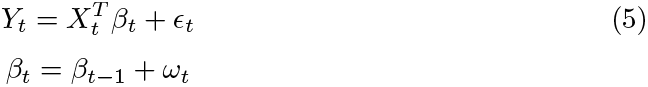

where

*Y*_*t*_ is the observed data **X**_*t*_ is a p-dimensional design vector of covariates an the interaction terms.

*β*_*t*_ is a p-dimensional time varying parameter vector (for both covariates an the interaction terms).

*ϵ*_*t*_ and *ω*_*t*_ are the observation and evolution error terms respectively where *ϵ*_*t*_ ∼𝒩 (0, *V*_*t*_) and *ω*_*t*_ ∼𝒩 (0, *W*_*t*_).

Since the formulated model is a multivariate Gaussian state space model, basing on the standard results about a multivariate Gaussian distribution pertaining its marginal and conditional distributions, the random vector of parameters *β*_*t*_ too has a Gaussian distribution and the same applies to the other respective marginal and conditional distributions.

Therefore, since all the relevant distributions are Gaussian, we use the means and variances to estimate the parameters (posterior means) through the following procedure:

i. Obtaining the prior distribution (basing on the evolution equation reflected in the second line of (3)). Considering the fact that a DLM is a Gaussian time space model, a normal prior distribution was used for a p-dimensional state vector, first obtaining the initial values where 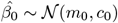 Attaining a one step ahead of the prediction of the parameter vector distribution 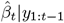 Obtaining the mean

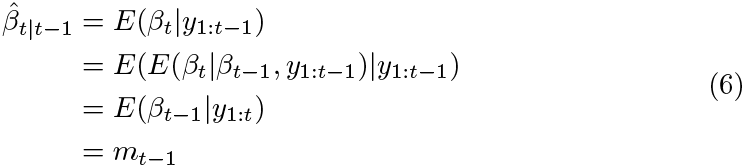 Also obtaining the predicted covariance matrix *c*_*t*|*t−*1_

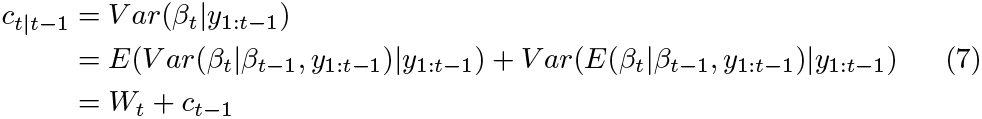 Therefore, 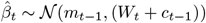 since our prior has been derived basing on a normal distribution, we can write its probability density function as follows:

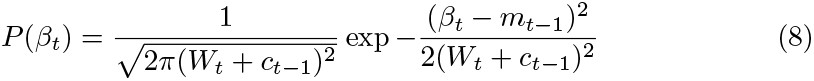
ii. Obtaining the likelihood distribution function (basing on the observation equation reflected in the first line of (3)) . Obtaining the predicted (one step) distribution function of *Y*_*t*_ (the observed value): 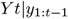 Let 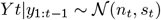 Obtaining the mean, *n*_*t*_

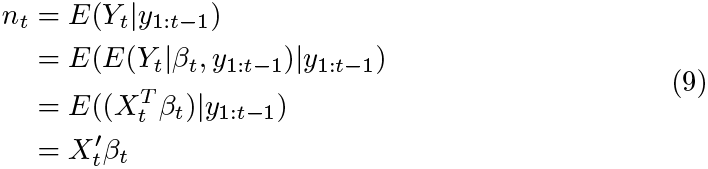 While

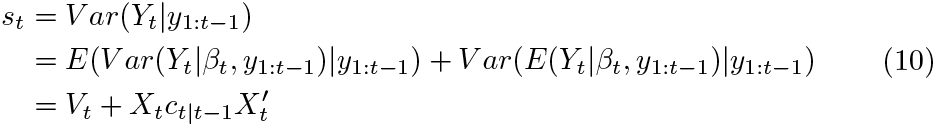 Therefore, 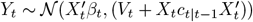 Since the observed value variable follows a normal distribution, we can write its likelihood function probability density function as follows;

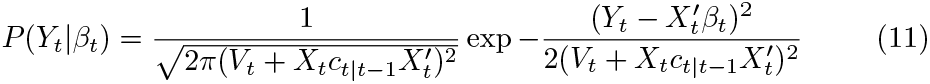
iii. Obtaining the posterior distribution From (4), expression derived from Bayes theorem, we are able to obtain the posterior probability density function by multiplying (11) with (8) and hence obtaining the probability density function of the posterior distribution function as follows;

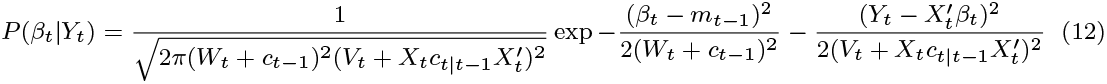 Since both the prior and likelihood distributions are guassian, then the resulting posterior is also guassian. If we let the posterior *β*_*t*_|*Y*_*t*_ *∼* 𝒩 (*d*_*t*_, *p*_*t*_), from the first property of a gaussian distribution pertaining the probability density function, we can be able to obtain the values of *d*_*t*_, *p*_*t*_ by simplifying the right hand side of (12);

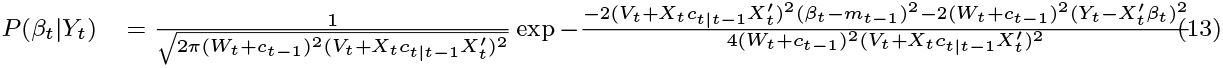

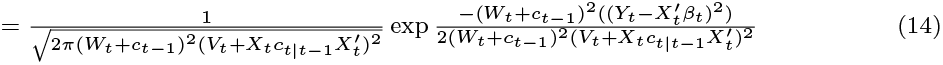

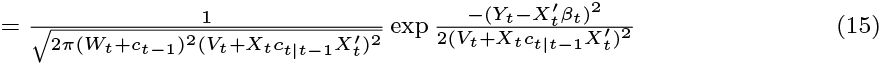

from (13),

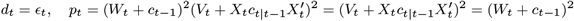 Therefor, 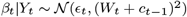

### Model Validation

To check the model fitness, analysis of residuals was done. Posterior means and credible intervals for parameters were estimated and by use of trace and means plots, convergence was tested.

To validate the model, the study conducted the following residual tests on; autocorrelation, heteroscedasticity, and normality using the Durbin-Watson test, the Breusch Pagan test, and the Shapiro-Wilk test respectively and hypotheses testing was done.

To verify the above model properties, tests were carried out using the following hypotheses:

*H*_01_: Residuals are not auto-correlated Vs *H*_11_: Residuals are auto-correlated.

*H*_02_: Residuals have constant variance Vs *H*_12_: Residuals do not have constant variance.

*H*_03_: Residuals are normally distributed *H*_13_ Vs *H*_01_: Residuals are not normally distributed.

### Test Statistics, Critical Values, and the Decision Rule

While testing for autocorrelation of residuals, the Durbin-Watson test was used. To obtain its test statistic, the following formula was used:

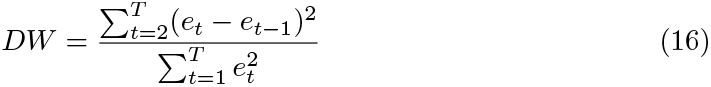

where

*e*_*t*_ = the residuals from the regression model.

T = the number of observations.

Pertaining the critical value and the decision rule, we based on the fact that the DW (value) ranges between 0 and 4; since the DW value approaching 2 indicates no autocorrelation, the value approaching 0 indicates positive autocorrelation, and the value approaching 4 indicates negative autocorrelation.

While testing for homoscedasticity of residuals, the Breusch Pagan test was used whereby to obtain its test statistic, the following formula was used:

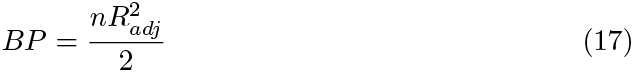

where

n = number of observations

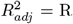 -squared value obtained by regressing the squared residuals from the original regression on the independent variables, their squares, and their cross products. This regression is known as Auxiliary regression.

About the critical value and the decision rule, the fact that the test statistic of Breusch Pagan follows a chi-square distribution with degrees of freedom equal to the number of independent variables in the auxiliary regression, the study compares the test statistic value with the critical (tabulated) value to decide on rejecting the null hypothesis.

While testing whether the residuals are normally distributed, Shapiro-Wilk’s test was used whereby the following formula was used to obtain its test statistic denoted as W:

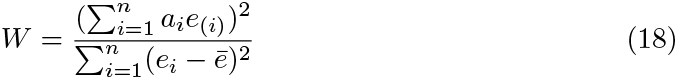

where

*e*_(*i*)_ = ordered residuals from smallest to largest.

*ē* = mean of the residuals.

*a*_*i*_ = constants derived from the expected values of the order statistics of a standard normal distribution computed using statistical software.

n = number of observations.

On the critical value and the decision rule, these were determined based on the fact that W ranges between 0 and 1; values of W close to 1 suggest that the residuals are approximately normally distributed whereas lower values indicate deviations from normality.

### Software and Tools

The study used R with the most relevant libraries for data processing and statistical data analysis. For instance the study used thr r-JAGS package while fitting the model.

## Results and Discussions

### Model Validation Results and Discussion

The following conclusions have been made regarding Model validation:

Basing on the results in output 1:

While testing for auto-correlation of residuals whereby the Durbin-Watson test was used, basing on the fact that the Durbin-Watson value (DW = 0.07253) ranges between 0 and 2, this indicates no auto-correlation of the residuals.

While testing for homoscedasticity of residuals under which the Breusch Pagan test was used, by the fact that the probability value of the of Breusch Pagan test statistic (p-value = 0.3755) is greater than 0.05, we fail to reject the null hypothesis and conclude that Residuals have constant variance.

While testing whether the residuals are normally distributed whereby Shapiro-Wilk’s test was used, basing on the fact that Shapiro-Wilk’s test value W ranges between 0 and 1; since W (W = 0.9963) is close or equal to 1, it suggests that the residuals are normally distributed.

Therefore, the model fulfills the expected assumptions of no autocorrelation, homoscedasticity and normality of residuals.

#### Listing 1.

Model Validation Test Statistics Durbin Watson Test

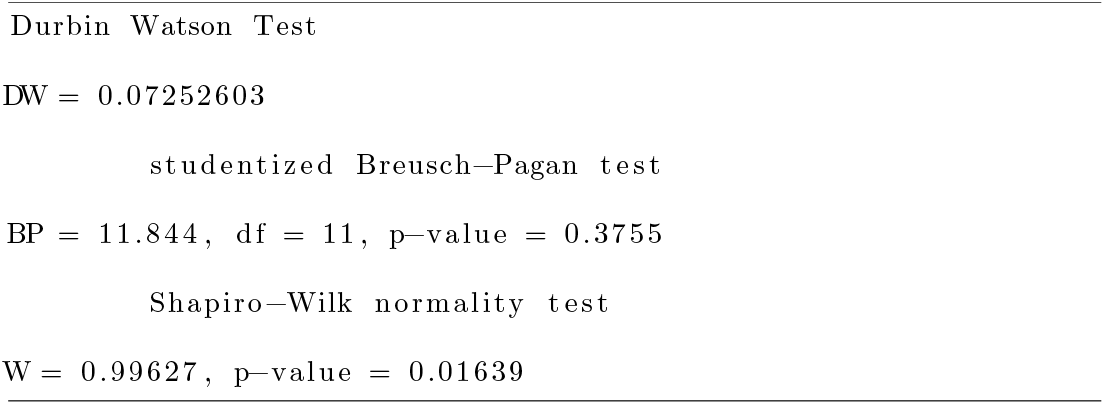

### Model Convergence Results and Discussion

From the sample of Traces and Density plots in the output in figures 1 and 2 attached, there is evidence that the model parameters are time dependent and a few are influenced by external factors as evidenced from the traces plots of variables and their density plots which too further prove that they follow a normal distribution.

**Fig 1.**
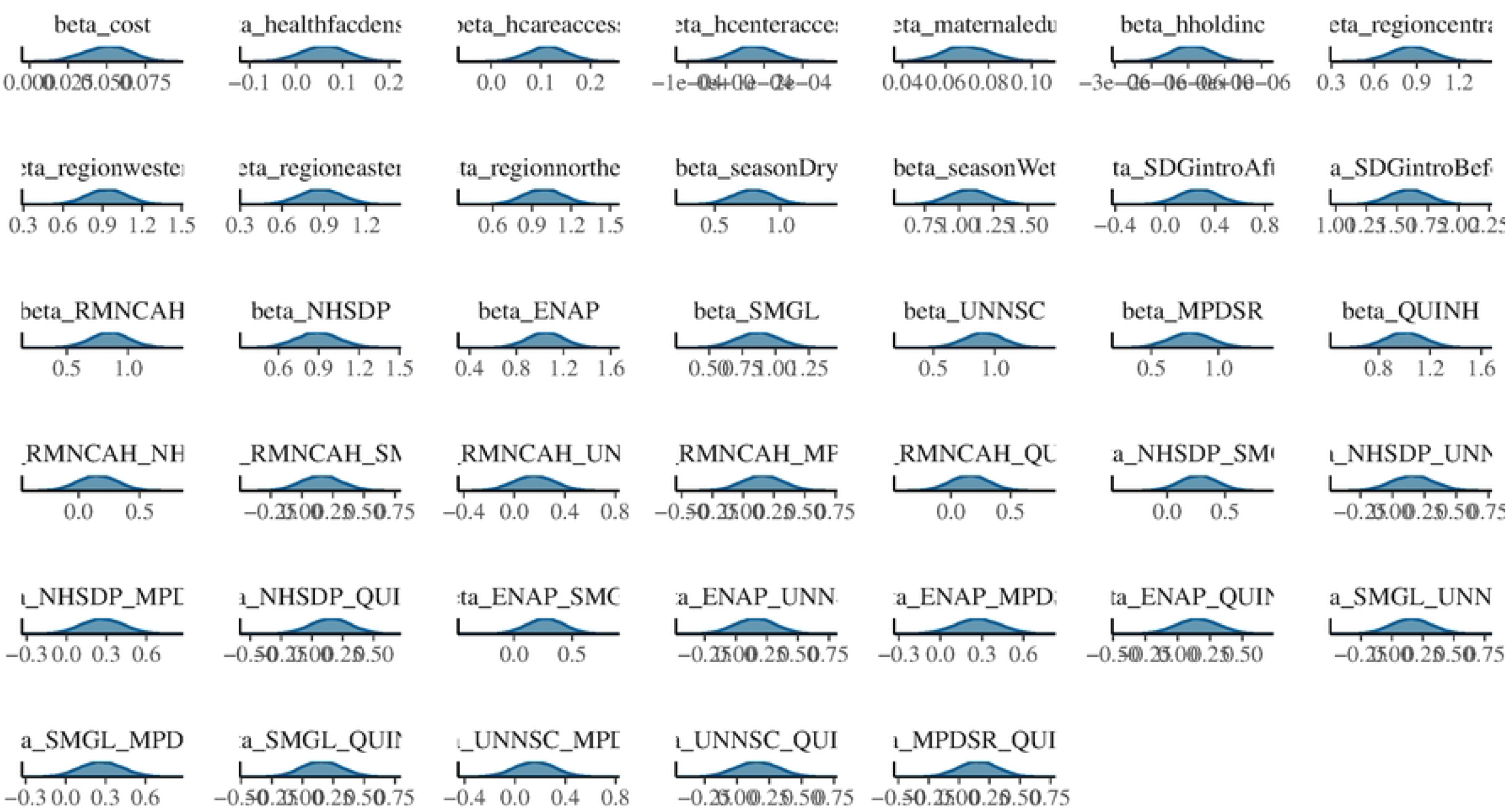
Trace plot of model parameters

**Fig 2.**
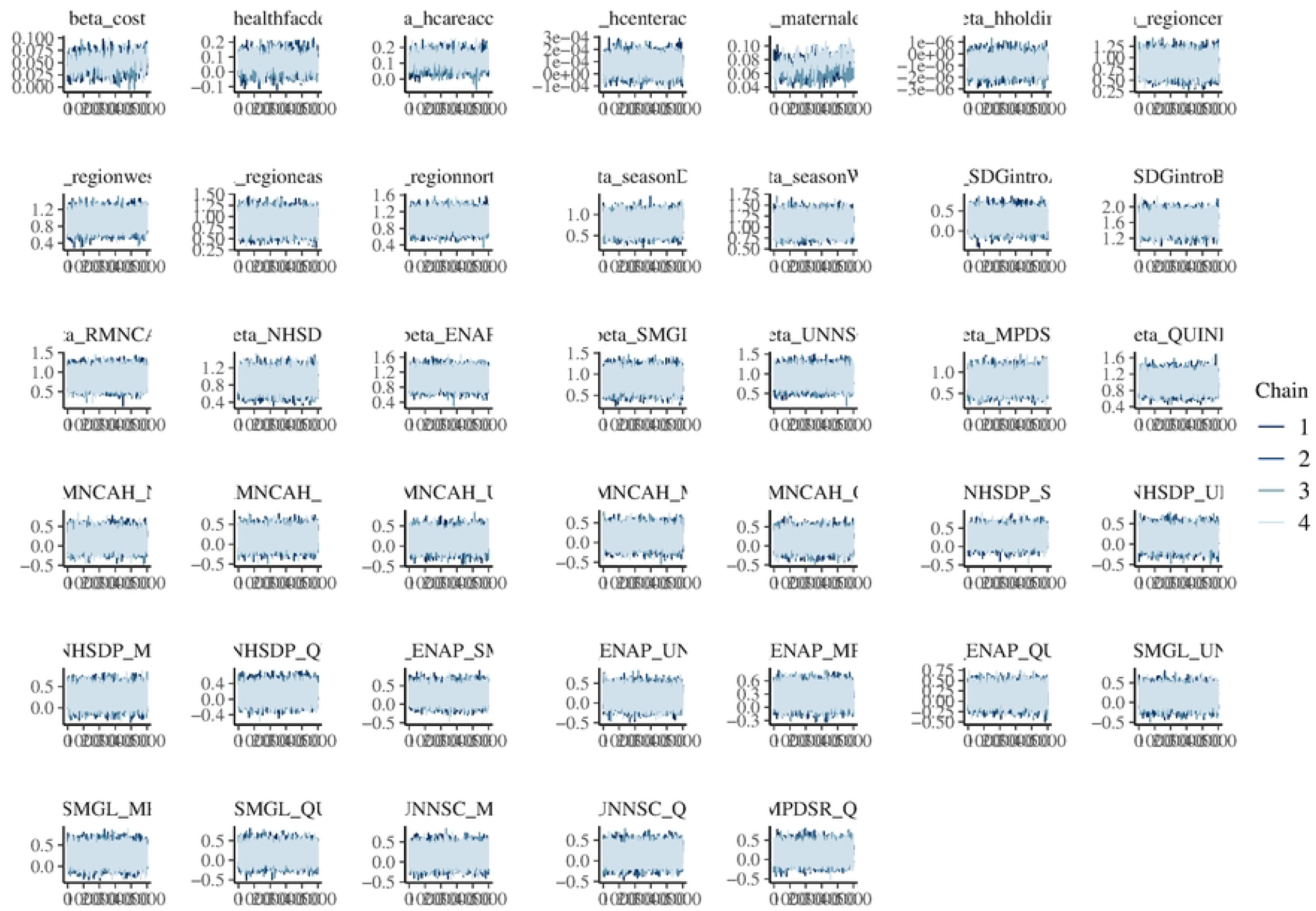
Density plot of model parameters

### Parameter Estimation Results and Discussion

From the means and quantiles of parameters as reported in 2, most of the variables have a positive influence on the dependent variable (Neonatal Mortality) with the exemption of health center access and household income as reported under means whereas regarding the quantiles, most of the variables significantly influence the outcome variables since their extreme quantiles are non zero.

#### Listing 2.

Means and Quantiles

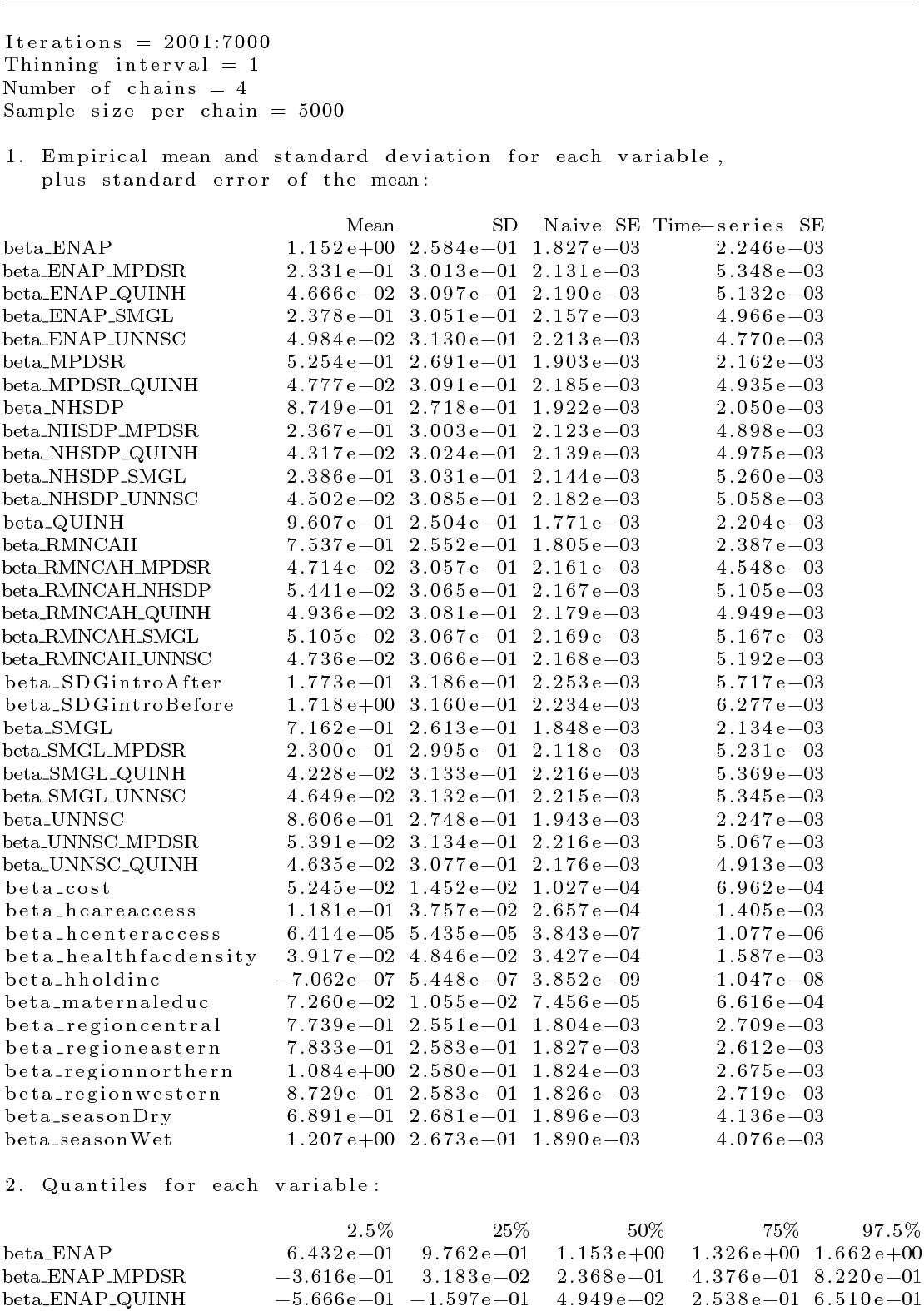
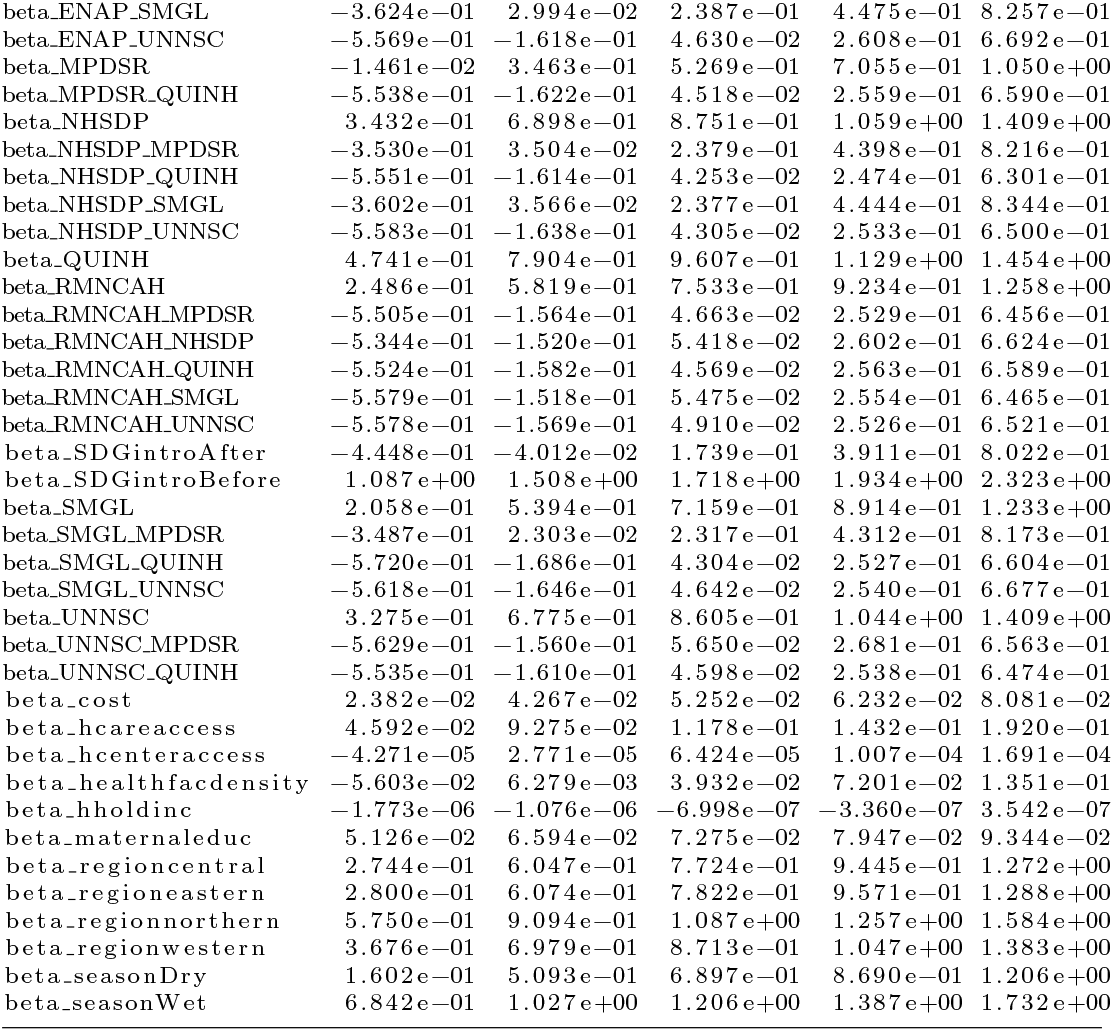

## Conclusion

The study has been able to develop a dynamic linear model that can be used to monitor health care policies individually and also assess the possibility of interaction between any two policies such wastage of resources on duplicated policies is avoided.

## Data Availability

All relevant data are within the manuscript and its Supporting Information files since the study relied on simulated data.

## Supporting information

**S1 Fig. Model Convergence (trace plot). Trace plot of model parameters**

**S2 Fig. Model Convergence (density plot). Density plot of model parameters**

**S1 File. R code** The R script used for data processing and analysis is provided as a supplementary file named ‘script.R’.

